# Molecular mechanisms promoting long-term cytopenia after BCMA CAR-T therapy in Multiple Myeloma

**DOI:** 10.1101/2024.05.22.24307750

**Authors:** Maria Luisa Palacios-Berraquero, Paula Rodriguez-Marquez, Maria Erendira Calleja-Cervantes, Nerea Berastegi, Aintzane Zabaleta, Leire Burgos, Diego Alignani, Patxi San Martin-Uriz, Amaia Vilas-Zornoza, Saray Rodriguez-Diaz, Susana Inoges, Ascensión Lopez-Diaz de Cerio, Sofia Huerga, Esteban Tamariz, Jose Rifón, Ana Alfonso-Pierola, Juan Jose Lasarte, Bruno Paiva, Mikel Hernaez, Paula Rodriguez-Otero, Jesus San Miguel, Teresa Ezponda, Juan Roberto Rodriguez-Madoz, Felipe Prosper

**Affiliations:** Hematology and Cell Therapy Department. Cancer Center Clinica Universidad de Navarra (CUN). IdiSNA; Hemato-Oncology Program. Cima Universidad de Navarra. IdiSNA; Computational Biology Program. Cima Universidad de Navarra. IdiSNA; Flow Cytometry Core. Cima Universidad de Navarra. IdiSNA; Immunology and Immunotherapy Department. Clinica Universidad de Navarra (CUN); Immunology and Immunotherapy Program. Cima Universidad de Navarra. IdiSNA; Data Science and Artificial Intelligence Institute (DATAI). Universidad de Navarra; Centro de Investigación Biomédica en Red de Cáncer (CIBERONC)

## Abstract

Hematological toxicity is a common side effect of CAR-T therapies, being particularly severe in relapsed/refractory multiple myeloma (MM) patients. In this study, we analyzed a cohort of 48 patients treated with BCMA CAR-T cells to characterize the kinetics of cytopenia, identify predictive factors and determine potential mechanism underlying these toxicities. The overall incidence of cytopenia was 95.74%, and grade>3 thrombocytopenia and neutropenia one month after infusion was observed in 57% and 53% of the patients and was still present after 1 year in 4 and 3 patients respectively. Presence of cytopenia at baseline and high peak inflammatory markers highly correlated with cytopenia persisting up to three months. To determine potential mechanisms underpinning cytopenias, we evaluated the paracrine effect of BCMA CAR-T cells on the differentiation of HSPCs using an *ex-vivo* myeloid differentiation model. Phenotypic analysis showed that supernatants from activated CAR-T cells (spCAR) halted HSPCs differentiation promoting more immature phenotypes, with reduced expression of granulocytic, monocytic and erythroid markers. Single-cell RNAseq demonstrated an upregulation of transcription factors associated with early stages of hematopoietic differentiation in the presence of spCAR (*GATA2, RUNX1* and *CEBPA)* and decreased activity of key regulons involved in neutrophil and monocytic maturation (*ID2* and *MAFB)*. Our results suggest that CAR-T cell activation negatively influences hematopoietic differentiation through paracrine effects inducing arrest of HSPCs maturation and contributes to the understanding of severe cytopenia observed after CAR-T cell treatment in MM patients. These results may identify regulatory mechanisms involved in alter hematopoiesis and could lead to alternative therapeutic strategies.

**KEY POINTS:**

- Long-lasting cytopenia after BCMA CAR-T therapy correlates with baseline cytopenia and peak inflammatory markers.
- Supernatants from activated BCMA CAR-T cells induced an inhibition of ex-vivo myeloid differentiation and rewiring of transcriptional programs associated with hematopoietic differentiation.

## INTRODUCTION

Chimeric antigen receptor (CAR)-T cell therapy has changed the treatment landscape for relapsed and refractory (R/R) B cell hematologic malignancies^1–3^. To date, triple-class exposed patients with R/R multiple myeloma (MM) present poor outcomes, with a median progression-free survival (PFS) of 3-4 months, and a median overall survival of 8-9 months^4,5^. Pivotal trials with BCMA-directed CAR-T cells have shown remarkable efficacy, achieving durable remissions ranging from 8-35 months PFS in this subgroup of patients with previously dismal prognosis^6,7^. Consequently, the US Food and Drug administration has approved two BCMA CAR-T cell products Idecabtagene Vicleucel (Ide-Cel)^8^ and Ciltacabtagene autoleucel (Cilta-Cel)^9^ for the treatment of R/R MM. Given the promising results observed with these treatments, other BCMA CAR-T cell products, as well as CAR-T cell therapies against other myeloma antigens (CD19, CD38, CD138, SLAMF7) are currently under evaluation^10^, trying to bring CAR-T cell therapies earlier in the management algorithm of the disease.

Adverse events associated with CAR-T cell therapy for R/R B-cell hematologic malignancies are common, with more than 80% of the patients developing toxicities due to the induction of immune effector cell responses^6,7,11^. Acute toxicities such as cytokine release syndrome (CRS) and immune effector-cell associated neurotoxicity syndrome (ICANS) have been extensively characterized from a physio-pathological and clinical standpoint^12–14^, and are thus well understood and managed^15,16^. However, mechanisms underpinning other toxicities such as hematologic toxicity and particularly long-term cytopenias, a frequent side effect of BCMA CAR-T therapy, are still poorly understood^17,18^. Initially attributed to lymphodepleting chemotherapy regimens, long-lasting cytopenias that are present more than 30 days post-infusion have been described across all CAR-T cell products, independent of their target antigen pointing towards a class effect that is independent of chemotherapy^19,20^. Recent studies focusing on CD19-directed CAR-T suggest a relationship between CRS and elevated inflammatory markers at baseline, and the development of long-lasting hematologic toxicity^21^. Data on BCMA CAR-T derived cytopenia comes from a few single-center retrospective descriptive analysis and points toward a similar relationship^22–24^.

In the current study, we characterized the kinetics of cytopenia in a cohort of patients with R/R MM receiving BCMA CAR-T cell treatment, evaluating their correlation with clinical and laboratory parameters. To delve into the mechanisms of these cytopenias, we studied the effect of activated CAR-T cell supernatant on *ex-vivo* hematopoietic differentiation. Our results using phenotypic and single cell transcriptional studies allowed us to identify abnormal transcriptional and regulatory programs involved in hematopoietic differentiation, providing molecular mechanistic insights driving prolonged cytopenia.

## MATERIALS AND METHODS

### Clinical metadata

Clinical metadata from 48 adult patients with R/R MM receiving BCMA CAR-T cell treatment were retrospectively analyzed. The study was approved by the Ethics Committee of the University of Navarra. Clinical data was collected after informed consent. We defined baseline values as those before lymphodepletion, with a latency period of up to 3 days for laboratory values, and 1 month for bone marrow infiltration, that was determined before apheresis and before the administration of any bridging therapy. Lymphodepletion was given according to each manufacturers’ instructions and was based on fludarabine and cyclophosphamide, followed by product infusion. Median follow-up was 10.5 months. Follow-up was continued until disease progression or death. Toxicity was graded according to Criteria for Adverse Events (CTCAE) v5.0. For long term hematological toxicity, we followed the criteria recently described by Rejeski et al, defining cytopenias after day +30 as long-term hematotoxicity^25^.

### Generation of activated BCMA CAR-T cell supernatants

A second-generation BCMA CAR construct was used^26^. Lentiviral vectors were produced in HEK293T cells following standard procedures^26^. CAR-T cells were generated from healthy donors as described^26^. Un-transduced T cells (UTD) and CAR-T cells were co-cultured separately with U266 cells at 1:1 ratio. Supernatants (spUTD and spCAR) were collected after 48h, cell debris were removed by centrifugation and clean supernatants were stored at -80°C until use.

### Cytokine quantification

Analysis of human cytokine levels in spCAR or spUTD supernatants was performed using a custom ProcartaPlex multiplex assay (Thermo Fisher Scientific) according to manufacturer’s instructions. 15-plex panel including IL-1β, IL-2, IL-4, IL-6, CD137, IL-8, IL-10, IL-17A, IFNγ, GM/CSF, TNFα, PERFORIN, IL-15, GRANZYME B and CD40L was used. Data was acquired in a Luminex™ 200™ Instrument System (Thermo Fisher Scientific).

### *Ex-vivo* myeloerythroid differentiation model

Mononuclear cells were obtained by Ficoll-Paque (GE Healthcare) density gradient centrifugation of bone marrow aspirates from healthy donors (n=3, range=18-22 y/o). All subjects provided written informed consent. HSPCs cells were stained using CD34-APC (clone 581; Beckman Coulter) and CD45-PerCPCy5.5 (clone HI30; Biolegend) and sorted in a BD FACSAria II (BD Biosciences). E*x-vivo* liquid culture differentiation assay was performed as previously described^27^. To evaluate the effect of supernatants from activated CAR-T cells, 2 mL of spCAR, spUTD or media as control were added to the differentiation process. OP9 cells, media, cytokines, and supernatants were renewed every 2-3 days.

### Flow cytometry analysis

Phenotypic characterization of neutrophilic, monocytic and erythroid lineages was performed after 24 days of differentiation. All antibodies were purchased from BD Biosciences unless otherwise stated (Table S1). Data was acquired on a BD FACSCanto II (BD Biosciences) and analyzed using the FlowJo Software version 10 (Tree Star).

### Single-cell RNA sequencing (scRNA-seq)

scRNA-seq was performed on cells cultured under spCAR and spUTD conditions after 24 days of differentiation using Chromium Single Cell 3′ Reagent Kit (10X Genomics) according to the manufacturer’s instructions. For spUTD samples, 12,463 cells were analyzed with an average sequencing depth of 26,678 reads/cell. For spCAR sample, 6,945 cells were analyzed with an average sequencing depth of 54,999 reads/cell. scRNA-seq data were demultiplexed, aligned to the human reference (GRCh38) and quantified using Cell Ranger (v6.0.1) from 10X Genomics. Further computational analysis was performed using Seurat (v3.1.5). Cells were filtered based on the number of detected genes, number of UMIs, and proportion of UMIs mapped to mitochondrial and ribosomal genes per cell. Using unsupervised clustering analysis with the resolution set to 0.6, a total of 15 cell clusters were identified. Afterwards, cell types and states were annotated using canonical marker genes as reference.

### Gene regulatory networks (GRN) analysis

Using the most variable 300 TFs and 3,000 genes, SimiC^28^ was run with the default parameters across spCAR and spUTD clusters. GRNs were plotted using the GRN incidence matrices provided by SimiC. The histograms for the different regulons were computed from the “regulon activity score” provided by SimiC. This score was also used to compute the regulatory dissimilarity score for the selected cell clusters.

### Statistical analysis for clinical metadata

Univariate analyses were performed via simple linear regression for variables on a continuous scale and by logarithmic regression for binary variables, using Spearman’s correlation. Mann-Whitney’s U test and Kruskall Wallis were used for categorical nonparametric variables. Statistical tests were two sided and significance was considered for p-values < 0.05. Statistical analyses were performed using IBM SPSS version 26.0 and GraphPad Prism version 9.3.1. The different tests used in this work are indicated in the figure legend.

## RESULTS

### BCMA CAR-T cell therapy in patients with relapse refractory myeloma is associated with long-lasting severe hematological toxicity

This study included 48 patients with a diagnosis of R/R MM treated with BCMA CAR-T cell therapy in our institution (Table 1). Patients were heavily pretreated with a median of 3 previous lines of treatment before apheresis (range 2-8), including 96% of patients with previous autologous stem cell transplantation (ASCT), and 21% with a history of two ASCT. No patient had received allogeneic transplant. 79% of patients were refractory to proteasome-inhibitors (PI), immunomodulatory agents (IMID), and anti-CD38 monoclonal antibodies (triple-class refractory), and 31% were penta-drug refractory (refractory to two PI, two IMIDs and anti-CD38 monoclonal antibodies). Only one patient had received prior BCMA directed treatment (Table S2). Of note, 29% of patients had extramedullary disease and 33% had high-risk cytogenetics.

**Table 1.** Patient characteristics.

| **Table 1. Patient characteristics** |  |
| --- | --- |
| **Characteristics** | **Number of patients** |
| Median age, y (range) | 57,5 (41-79) |
| Median number of previous lines of treatment (range) | 3 (2-8) |
| Previous ASCT | 46 (96) |
| >1 ASCT (%) | 10 (21) |
| **Disease characteristics** |  |
| High risk cytogenetics* | 16 (33) |
| Extramedullary disease | 14 (29) |
| Number of focal lesions |  |
| - None | 4 (8.3) |
| - 1-3 | 14 (29.2) |
| - 4-10 | 6 (12.5) |
| - >10 | 24 (50) |
| Number of lytic lesions |  |
| - None | 5 (10.4) |
| - 1-3 | 8 (16.7) |
| - 4-10 | 13 (27.1) |
| - >10 | 22 (45.8) |
| Triple-drug refractory | 38 (79) |
| Penta-drug refractory | 15 (31) |
| **Baseline blood count** |  |
| Median hemoglobin, g/dL (95% CI) | 11.2 (7.9-14.5) |
| Median platelet count per uL, (95% CI) | 139500 (34000-387000) |
| Median ANC per uL, (95% CI) | 2355 (2010-285) |
| Hemoglobin < 8g/dL (%) | 1 (2) |
| Platelets < 75.000/uL(%) | 4 (8) |
| ANC < 1000 cells/uL (%) | 3 (6) |
| **Median markers of inflammation at baseline (95% CI)** |  |
| Lactate dehydrogenase, U/L | 237 (213-275) |
| Ferritin,ng/mL | 368 (279-642.8) |
| D dimer, U/L | 605 (410-740) |
| Fibrinogen, mg/dL | 432 (364-506) |
| Patient baseline characteristics prior to CAR T cell infusion.<br>*High risk cytogenetics includes t(4;14), t(14;16), t(14;20), and/or del(17p). Triple-drug refractory disease indicates refractory to an IMiD, PI and daratumumab; penta-drug refractory indicates refractory to lenalidomide, pomalidomide, bortezomib, carfilzomib and daratumumab.<br>Abbreviations: ANC absolute neutrophil count; ASCT autologous stem cell transplant. |  |

The main adverse events observed after CAR-T cell therapy included CRS reported in 89.6% of patients (Table 2). Severe CRS events (grade ≥ 3) were seen in 12.5% of patients, and any grade ICANS was seen in 20.6% with grade ≥ 3 in 4% of patients. These complications occurred mostly in the first month after CAR-T cell treatment, and the median duration of these complications was 5.5 days (3.5-7.6, 95% CI) for CRS and 2.5 days (1.4-3.5, 95% CI) for ICANS. One patient in our cohort developed a delayed treatment related grade 3 parkinsonism syndrome^29^. The overall incidence of any grade cytopenia was 95.7%, with anemia, neutropenia and thrombocytopenia of any grade reported in 98%, 98% and 76% of the patients, respectively (Table 2). Cytopenia were seen across all BCMA CAR-T cell products and regardless of the number of prior lines of therapy. Median time to neutrophil (ANC >1,000/uL), platelet (>100,000/mm3) and hemoglobin (>9g/dL) recovery was 45, 90 and 90 days respectively. Regarding severity and duration of cytopenia one month after infusion, grade 2 anemia persisted in 17% of the patients, and more than half of patients had grade ≥ 3 neutropenia (53.1%) and thrombocytopenia (57.4%) (Fig. 1). The proportion of patients with grade ≥ 3 neutropenia and thrombocytopenia remained high after two months (30.95% and 40.5% respectively), three months (28% and 33.3%) and six months after infusion (21% and 21.4%). Febrile neutropenia was reported in 38% of patients, with grade ≥ 3 infections in the first month after treatment seen only in 10.4% of the patients (Table 2). One year after CAR-T cell infusion 4 patients still had platelet counts <50,000/mm^3^ and 3 patients had neutrophil counts <1,000/mm^3^. One patient presented a severe bleeding event in the central nervous system (grade ≥ 3).

**Table 2.** Main toxicities and management.

| **Table 2. Main toxicities and management** |  |
| --- | --- |
| **Toxicity** | **Value, N = 48, n (%)** |
| **Hematologic toxicity up to month 1 after infusion**<br>**Anemia**<br>- Any grade<br>- ≥ grade 3<br>**Thrombocytopenia**<br>- Any grade<br>- ≥ grade 3<br>**Neutropenia**<br>- Any grade<br>- ≥ Grade 3<br>- Profound (ANC < 100 cells/uL) | 47 (98)<br>30 (62.5)<br>37 (76)<br>30 (62.5)<br>47 (98)<br>47 (98)<br>12 (25) |
| **Supportive therapy for cytopenia**<br>G-CSF<br>TPO<br>EPO<br>CD34+ boost<br>pRBC transfusion < 30 days after infusion<br>pRBC transfusion > 30 days after infusion<br>Platelet transfusion < 30 days after infusion<br>Platelet transfusion > 30 days after infusion. | 39 (81.25)<br>4 (8.3)<br>8 (16.6)<br>1 (2)<br>33 (68.7)<br>6 (12)<br>25 (52)<br>8 (16.6) |
| **ICANS grade**<br>0<br>I<br>II<br>III<br>IV | 38 (79.1)<br>7 (14.6)<br>1 (2)<br>2 (4)<br>0 (0) |
| **CRS grade**<br>0<br>I<br>II<br>III<br>IV | 5 (10.4)<br>22 (45.8)<br>15 (31.3)<br>4 (8.3)<br>2 (4.2) |
| **Treatment for ICANS/CRS**<br>Steroids<br>Tocilizumab<br>Anakinra | 17 (35)<br>34 (70.8)<br>3 (6) |
| **Infection**<br>Any grade<br>Grade ≥ 3 | 18 (37.5)<br>5 (10.4) |
| Main toxicities and management after CAR T cell therapy.<br>Abbreviations: ANC absolute neutrophil count; CRS cytokine release syndrome; ICANS immune effector-cell associated neurotoxicity syndrome; G-CSF granulocyte colonies stimulating factor; EPO erythropoietin; pRBC packed red blood cells; TPO thrombopoietin analogues. |  |

**Figure 1.**
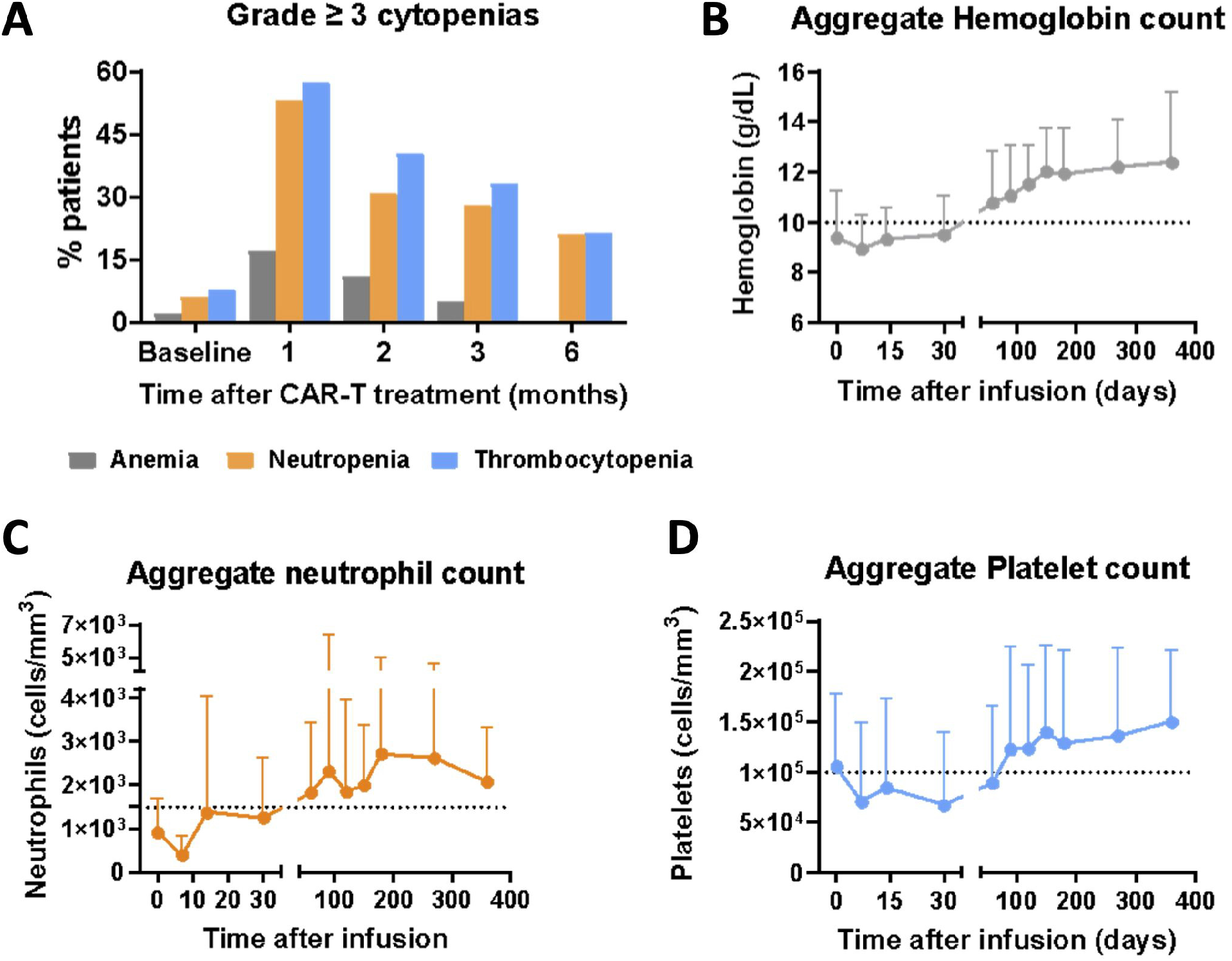
Long-lasting severe cytopenia was developed in patients treated with BCMA CAR-T cell therapy. **(A)** Prevalence of grade ≥ 3 anemia, thrombocytopenia, and neutropenia at baseline (0) and 1, 2, 3 and 6 months after infusion of CAR-T cell treatment. Evolution of aggregate hemoglobin **(B)**, neutrophil **(C)** and platelet **(D)** counts from CAR-T infusion up to one year of follow up.

Patients received supportive therapy with transfusion of packed red blood cell in 68% of patients in the first month after CAR-T cell infusion and 52% received pooled platelet transfusion, with 12% and 16.3% requiring transfusion in the following 3 months. G-CSF was administered in 89% of patients, mainly in the first month after treatment. In 17% of the patients, a recovery of neutrophil counts associated with G-CSF treatment was observed before day 28 with a subsequent decrease after G-CSF discontinuation. In 15% of the patients G-CSF administration was required in the first 3 months after CAR-T therapy. EPO and TPO were used in 16.6 and 8.3% of patients, and these treatments were started more than one month after CAR-T cell treatment in all cases (Table 2). These results indicate that hematological toxicity is a common and long-lasting toxicity associated with BCMA CAR-T cells.

### Baseline cytopenia, inflammatory markers and high-risk cytogenetics correlate with prolonged cytopenia

To define potential factors associated with hematological toxicity, we analyzed the relation between cytopenia and clinical and laboratory findings. Baseline hemoglobin level and thrombocytopenia were the best predictive biomarkers of long lasting cytopenia (Fig. S1). Thus, lower baseline hemoglobin levels correlated with lower hemoglobin, platelet, and neutrophil counts at 1, 3 and 6 months after infusion (Table S3). Similarly, baseline thrombocytopenia correlated with lower hemoglobin, platelet and neutrophil counts 3 months after infusion. Moreover, patients with high-risk cytogenetics did also significantly correlate with lower hemoglobin, platelet, and neutrophil counts 1 and 2 months after infusion. However, baseline bone marrow infiltration before CAR-T cell treatment did not correlate with later development of hematologic toxicity or its duration. Other disease-related factors (prior number of treatments, triple or penta-drug exposure, extramedullary disease, number of focal and lytic lesions, response to bridging therapy, use of alkylants in bridging therapy) and patient related factors (age, ECOG, baseline renal function, D dimer) were not associated with cytopenia either.

Regarding inflammatory markers, high baseline ferritin correlated with lower hemoglobin levels up to 6 months post infusion and lower platelet counts up to 5 months after infusion, but not with neutropenia (Table S3). Similarly, peak ferritin levels 3 days after infusion of CAR-T products correlated with lower blood counts at 1 month (Table S3). Interestingly, persistent high ferritin levels 1 month after infusion correlated with lower hemoglobin and platelet counts 2 and 3 months after infusion, indicating that high ferritin levels, marker of sustained inflammation, are associated with long lasting cytopenia. This might suggest that CAR-T cell-driven inflammation could be responsible for the hematopoietic toxicity observed after BCMA CAR-T cell therapy. To corroborate the paracrine effect of CAR-T cells on HSPCs further research into molecular mechanisms was conducted.

### Delayed *ex vivo* HSPC differentiation is mediated by paracrine effect of CAR-T cells

Except for B-cell progenitors and plasma cells^30^, HSPCs do not express the BCMA antigen and a direct effect of CAR-T cells on hematopoiesis is unlikely. Thus, to test a putative CAR-T-driven paracrine effect we used a previously described *ex-vivo* liquid culture differentiation assay^24^ where differentiation of CD34^+^ HSPCs was induced in the presence of supernatants from activated BCMA CAR-T cells (spCAR) or untransduced T cells (spUTD), as control (Fig. 2A). The presence of 15 different cytokines related to T cell activation, T cell effector function, such as IFNγ, TNFα, IL2 and IL6, as well as T cell polarization (Th response), were measured in the supernatants and showed significantly higher cytokine levels in spCAR in comparison with spUTD (Fig. 2B and S2). Upon 12 days of cell culture differentiation, no differences were detected in the number of cells among the 3 main differentiation lineages (neutrophilic, monocytic and erythroid) (Fig. 2C and S3). However, after 24 days, the number of CD10^-^ neutrophil precursors was significantly reduced in spUTD in comparison with spCAR condition. Moreover, cells cultured in the presence of spCAR showed significant decreased numbers of more differentiated cells in comparison with spUTD in all three lineages, neutrophilic (CD10^+^CD16^+^), monocytic (CD14^+^CD35^+^) and erythroid (CD71^+^CD36^+^) (Fig. 2C-D and S3). These data suggest that strong proinflammatory conditions produced by activation of CAR-T cells significantly affects hematopoiesis, potentially contributing to long-term cytopenia in patients undergoing BCMA CAR-T therapy.

**Figure 2.**
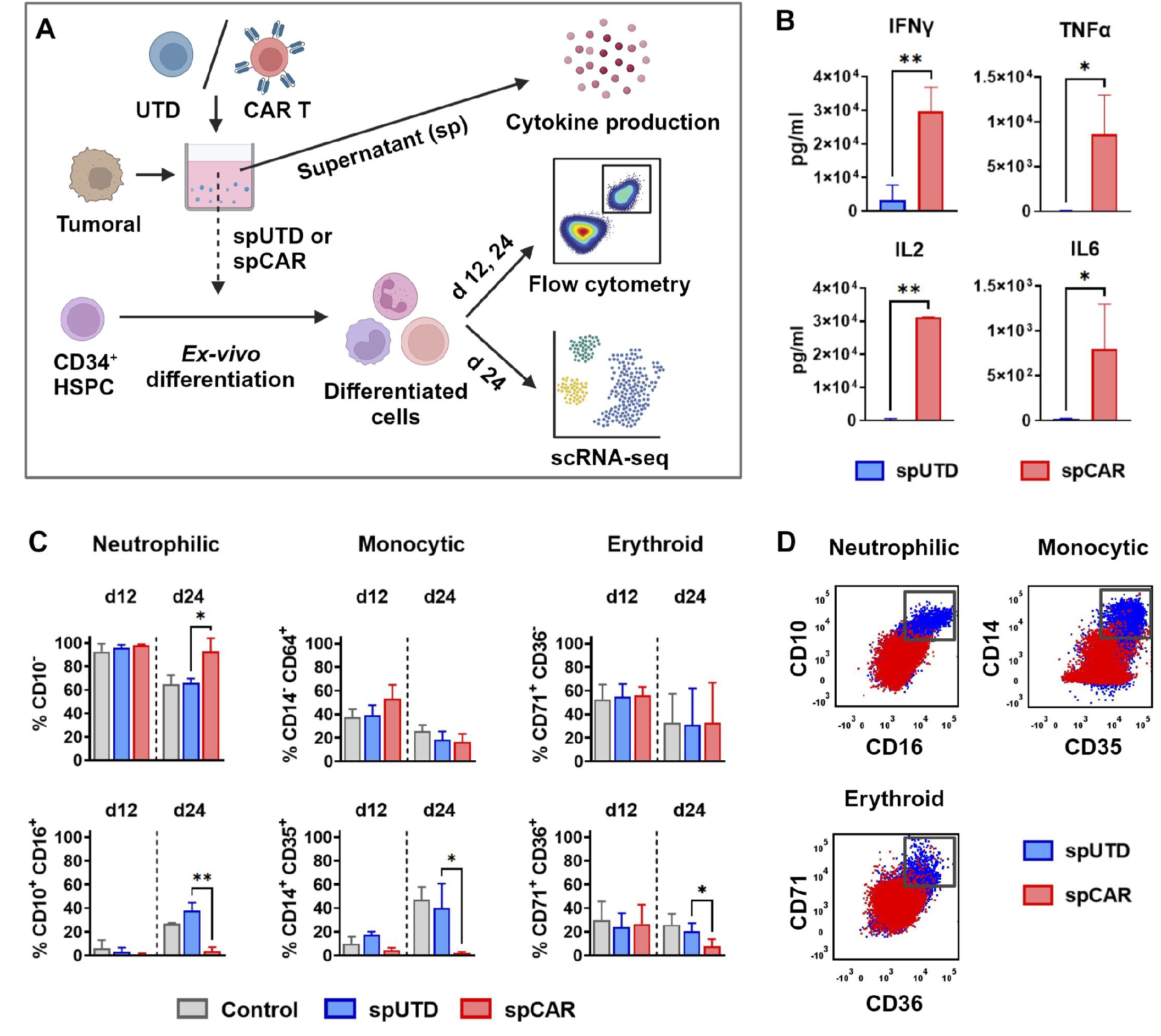
HSPCs differentiated with the supernatant of activated CAR-T cells presented less mature phenotypes. **(A)** Schematic representation of the *ex-vivo* myeloerythroid differentiation model employed. CD34^+^ HSPCs were harvested and subjected to differentiation under three conditions: addition of supernatant produced by the coculture of untransduced lymphocytes (spUTD) or BCMA CAR-T cells (spCAR) with MM tumoral cell line U266 for 48h, and control condition without addition of supernatant. Cytokine production was measured in the supernatant. The phenotype obtained after 12 and 24 days of *ex-vivo* differentiation of the HSPCs was studied by next generation flow cytometry. scRNA-seq was performed after 24 days of *ex-vivo* differentiation. **(B)** Concentration of IFNγ, TNFα, IL2 and IL6 cytokines in the supernatant of UTD (blue) and activated BCMA CAR-T cells (red) after 48h of co-culture with MM cell line U266 at 1:1 effector:target ratio. **(C)** Proportion of HSPCs differentiated in the control (gray), spUTD (blue) and spCAR (red) conditions. Analysis of less differentiated (upper panel) and more differentiated (lower panel) cells is shown for the three lineages, neutrophilic (CD10^-^; CD10^+^CD16^+^), monocytic (CD14^-^CD64^+^; CD14^+^CD35^+^) and erythroid (CD71^+^CD36^-^; CD71^+^CD36^+^). The proportion of cells achieving mature myeloerythroid phenotypes was significantly lower in the spCAR group. **(D)** FACS gating results of HSPCs differentiated under spUTD (blue) or spCAR (red) conditions at day 24 of differentiation. Gates of more differentiated cells are shown for neutrophilic (CD10^+^CD16^+^), monocytic (CD14^+^CD35^+^) and erythroid (CD71^+^CD36^+^) lineages respectively. Welch’s test (B) and unpaired t-test (C) were used. *p<0.05; **p>0.01.

### Single-cell transcriptional characterization of abnormal hematopoietic differentiation

To further understand the effect of supernatants from activated BCMA CAR-T cells on hematopoiesis, we performed scRNA-seq on CD34^+^ HSPC cultured for 24 days in the presence of spCAR or spUTD. A total of 14,248 cells were integrated (4,639 from spCAR and 9,609 from spUTD), and 15 clusters were defined (Fig. S4A-B). Clusters containing high levels of ribosomal genes and those in phase G2M/S of cell cycle were removed (Fig. S4C), and the rest were annotated based on the expression of canonical markers^31–35^. While clusters 0, 5, 10, and 13 were composed mainly of cells from spCAR, clusters 1, 3, 4, 11, and 12 included cells from spUTD (Fig. S4D). Clusters enriched in spUTD cells showed transcriptional profiles of mature granulocytes, monocytes, or macrophages. In contrast, clusters mainly composed of cells from spCAR, corresponded to cell populations with immature transcriptomic profiles, with enrichment in transcription factors (TFs) and genes involved in early hematopoietic homeostasis (Fig. 3A-B). To gain further insights into the potential transcriptional mechanisms involved in the abnormal hematopoietic differentiation we examined the main 3 trajectories. Consistent with immunophenotypic results, the transcriptional profile of cells cultured with spUTD was enriched in signatures of mature neutrophils, with expression of *PRTN3, ELANE, MPO, LTF, MMP8*, described as canonical neutrophils markers^36–40^ (Fig. 3C). In contrast, cells cultured with spCAR were enriched in *RUNX1*^41,42^ and *GATA2*^43–45^, TFs present in early hematopoietic precursors. These cells also presented higher expression of *CXCR4*, a TF of high relevance in HSPC and early neutrophil trafficking from the bone marrow^40,46,47^,and *CEBPA*, a TF largely involved in neutrophil differentiation with important role in myeloid priming^48^ (Fig. 3C). Moreover, mature monocytes within cells exposed to spUTD expressed *CD14*, a surface marker antigen of monocyte/macrophage lineage, as well as *CXCL16, IL18, CD74*^32,33,49^. In contrast, cells exposed to spCAR expressed *VSTM1, CLU, PADI4, CCL23*^32,34,50^ markers associated with immature monocytes (Fig. 3D). We also observed erythroid precursors within cells cultured with spCAR that presented differential expression of *RHEX, TPSAB1* and *KIT*^51–53^ (Fig. 3E). Other mature populations, such as antigen-presenting cells characterized by the expression of *HLA* genes and *CD1* family genes (*CD1A, CD1D*), were only observed in HSPCs cultured with spUTD (Fig. 3B). In line with these results, gene ontology (GO) analysis revealed that HSPCs exposed to spUTD presented enrichment in pathways related with granulocyte chemotaxis, neutrophil chemotaxis, migration, degranulation, and transcriptomic profile associated with macrophage features, highlighting the mature phenotype of neutrophilic and monocytic populations (Fig. 3F and S4E). On the other hand, HSPCs exposed to spCAR were enriched in pathways involved in the regulation of HSPC differentiation processes (Fig. 3G and S4F). Moreover, neutrophil, monocyte, and erythroid precursors presented increased apoptotic-related genes (Fig. S4G), with erythroid progenitors also showing negative regulation of G2/M and mitotic cell cycle phase transition, suggesting a decreased proliferation rate (Fig. 3G). Collectively, these data suggest a paracrine effect of CAR-T cells that leads to a more immature phenotype of HSPC differentiated in the presence of spCAR, increased apoptotic rate, and diminished proliferative activity of these cells, potentially explaining the long term cytopenias observed in these patients.

**Figure 3.**
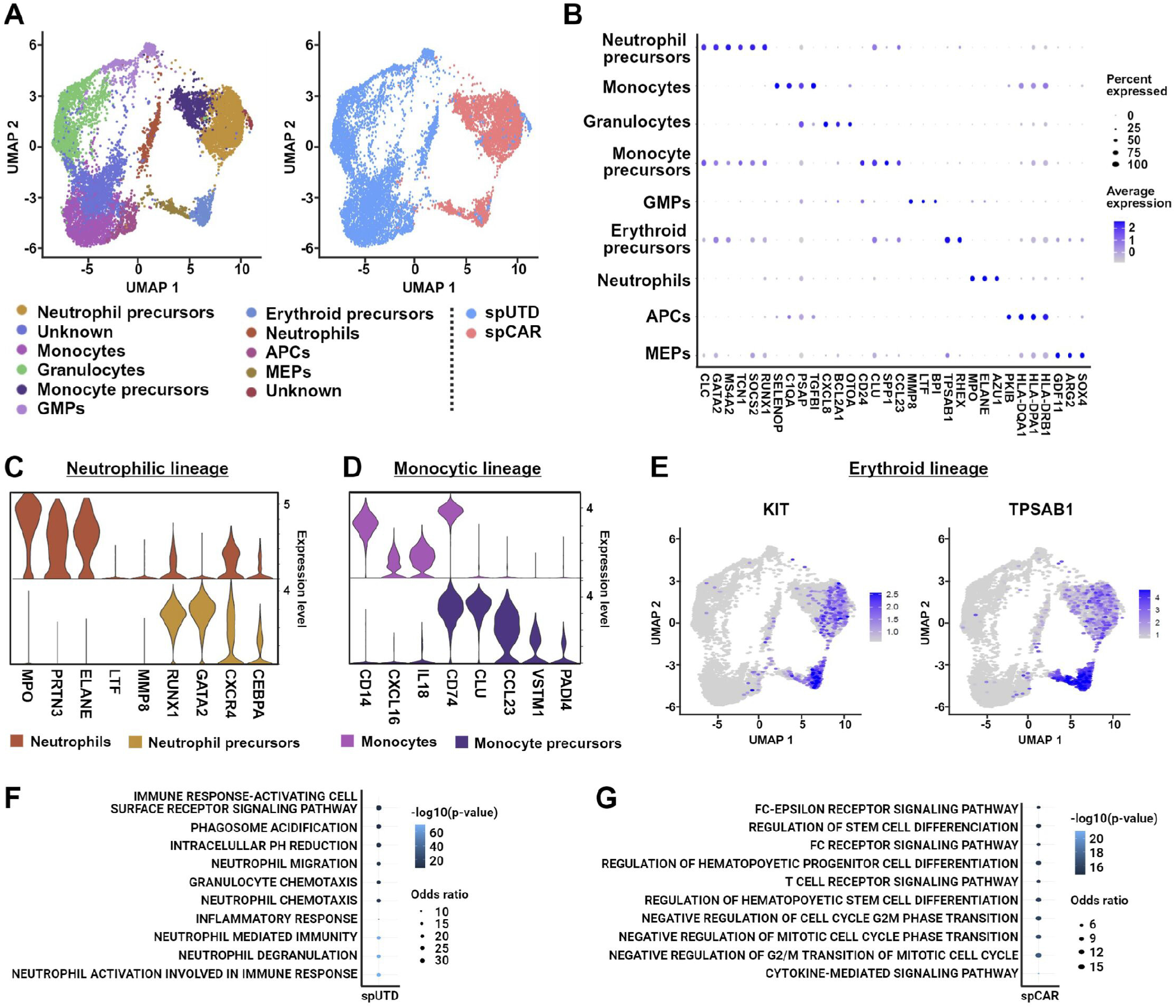
Characterization of differentiated CD34^+^ cells at single cell level. scRNAseq of *ex-vivo* liquid culture differentiation samples of healthy CD34^+^ cells at day 24 after addition of spCAR or spUTD as control was performed. **(A)** An overview of the 8259 cells that passed QC and filtering for subsequent analyses in this study. On the left, UMAP plot showing the 11 clusters that were analyzed and annotated On the right, UMAP plot showing the distribution of cells from each condition (spUTD or spCAR). **(B)** Dotplot with the expression of canonical markers. **(C)** Violin plots of cell markers for mature neutrophils (*MPO, PRTN3, ELANE, LTF*, and *MMP8*) and precursors of neutrophils (*RUNX1, GATA2, CXCR4*, and *CEBPA*). **(D)** Violin plots of cell markers for mature monocytes (*CD14, CXCL16, IL18*, and *CD74*) and monocyte precursors (*CLU, CCL23, VSTM1, and PADI4*). **(E)** UMAP plot showing the expression of KIT and TPSAB1, which are mainly distributed within erythroid precursors. Finally, gene ontology analysis of granulocytes **(F)** and erythroid precursor **(G)** clusters, corresponding to cells exposed to spUTD and spCAR respectively, showed pathways that confirm these phenotypes.

### Differential GRNs are seen in neutrophil and monocyte lineages exposed to activated CAR-T supernatants

To elucidate how supernatants from activated CAR-T alter GRNs, we applied SimiC^28^, observing regulons differentially activated between HSPCs cultured in the presence of spCAR or control (Fig. S5A). Cells exposed to spCAR presented increased activity of *JUND, JUNB* or *FOS*^54^ regulons, which are associated with myeloid differentiation with an increase in immature progenitors (Fig. S5B). We next compared the GRN activity of precursors from each lineage identifying a high dissimilarity score in the neutrophilic and monocytic lineage (Fig. 4A). In particular, increased activity of regulons *ID2* and *CEBPD* in neutrophils corresponded with their mature phenotype, as they are key TFs of terminally differentiated neutrophils^55,56^ (Fig. S5C). In contrast, the increased activity in neutrophil precursors of regulons *KFL6* and *CEBPB*, key TFs for neutrophil differentiation from myeloid precursors ^57,58^, explained the immature phenotype of these cells treated with spCAR (Fig. 4B). Similarly, in the monocytic lineage, increased activity of regulons involved in myeloid differentiation, together with decreased activity of *MEF2C* or *MAFB*^59,60^ observed in the presence of spCAR, explained their less differentiated phenotype (Fig. 4B).

**Figure 4.**
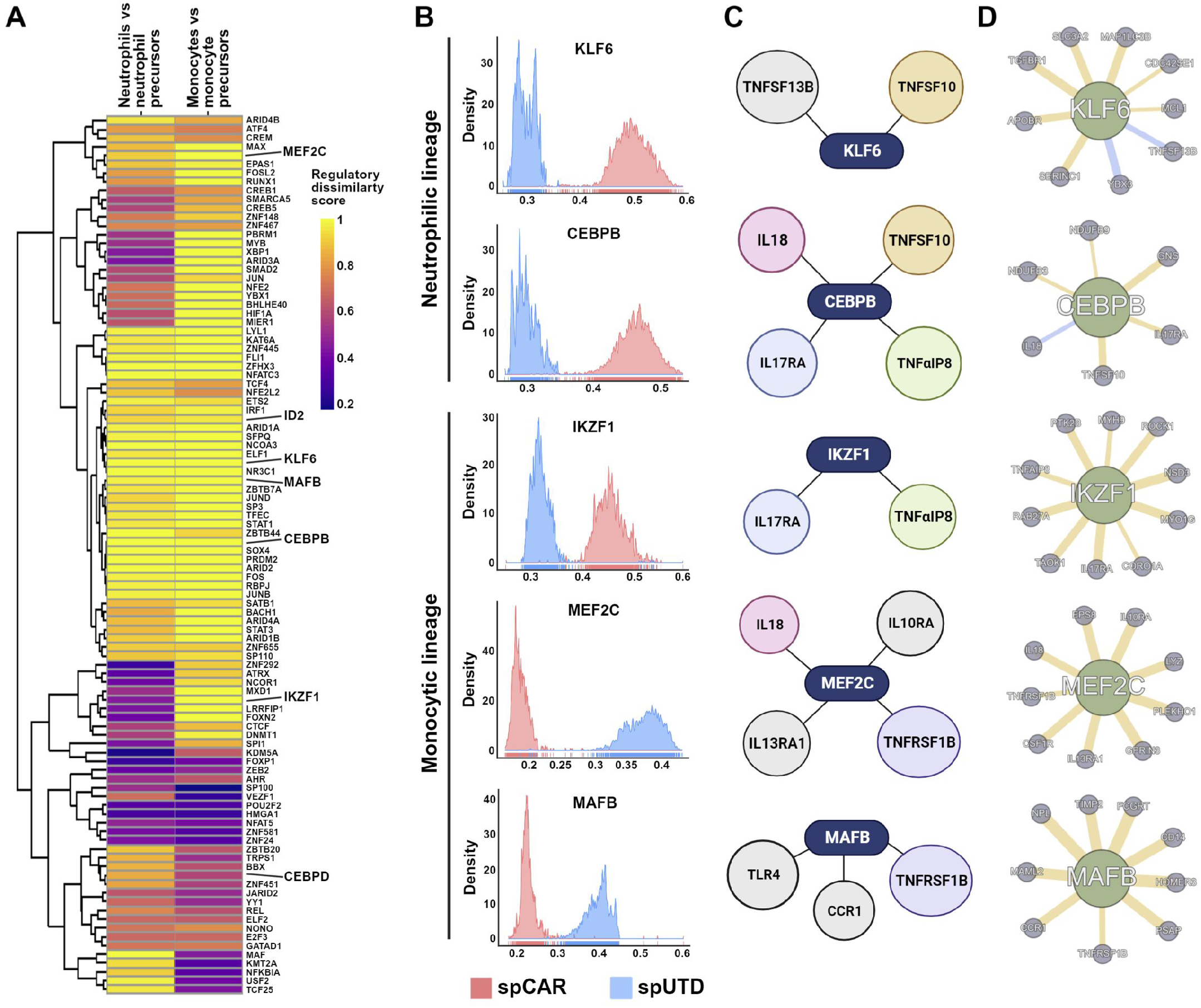
Analysis of GRNs in cells differentiated in presence of spCAR or spUTD. SimiC was applied to infer GRNs associated to more mature phenotypes or still in differentiation process. **(A)** Heatmap showing the regulatory dissimilarity score between cells exposed to spUTD or spCAR of the different regulons within neutrophil lineage cells and monocytic lineage cells. **(B)** Histograms showing the activity score of *KLF6* and *CEBPB* regulons, which are associated to neutrophilic lineage, and *IKZF1, MEF2C*, and *MAFB* regulons, associated to monocytic lineage between cells exposed to spUTD or spCAR. **(C)** Representative scheme of cytokine-related genes that belong to the regulons *KLF6, CEBPB, IKZF1, MEF2C*, and *MAFB*. **(D)** Networks of the transcription factors *KLF6, CEBPB, IKZF1, MEF2C*, and *MAFB* and their top related genes.

A detailed analysis of the TFs and their associated genes revealed genes related to proinflammatory cytokines, such as *IL18*, cytokine receptors, such as *IL17RA*, and members of TNF superfamily among *KFL6, CEBPB*, and *IKZFI* regulons, highlighting the importance that the inflammatory environment caused by the supernatant from activated CAR-T cells may have on HSPC (Fig. 4C-D). Moreover, *MEF2C* and *MAFB* regulons were also associated with genes related to cytokines, chemokines, or their receptors. Since IL13 regulates monocytic function^61^, the presence of its receptor among genes related to *MEF2C* regulon suggested the role of this cytokine in the maturation of these cells cultured with spUTD (Fig. 4C-D). Overall, our GRN analysis provides mechanistic insights into the regulatory networks underlying the phenotypic differences observed in HSPC differentiation in the presence of supernatants from activated CAR-T cells. All these data support our hypothesis that activation of CAR-T induces a paracrine effect that affects HSPC differentiation, resulting in long-term cytopenia in patients undergoing BCMA CAR-T therapy.

## DISCUSSION

Hematologic toxicity is one of the most frequent adverse events derived from CAR-T therapies and, not surprinsingly, in the current study cytopenia emerged as the most frequent toxicity after BCMA CAR-T cell treatment. Previous clinical studies, leading to the approval of Ide-Cel^2,8^ and Cilta-Cel^3,9^, showed high incidence of grade 3 and 4 cytopenia (neutropenia 89%, thrombocytopenia 52% and anemia 60%)^1–3,9^, a frequency similar to that found in our study. A recent report including a small cohort of patients receiving BCMA and CD19 CAR-T cell therapy for R/R MM focusing on hematologic toxicity^24^ reported prolonged cytopenia (defined as grade 3 or higher after day 28) in 58% of the patients. In our study the median time to recovery was remarkably higher, with over half of patients exhibiting grade ≥3 neutropenia (53%) and thrombopenia (57%) one month after infusion. Of note, one fifth of patients presented persistent severe thrombocytopenia and neutropenia six months after infusion (20% and 21%, respectively). The incidence of other CAR-T cell derived toxicities, CRS and ICANS was similar to that seen in KarMMa and CARTITUDE studies, although duration of symptoms was shorter probably in relation to a higher use of tocilizumab (79%) and steroids (35%) in our series. In this sense, even though in our series we did not find association between tocilizumab administration and cytopenia, its use has been reported to induce neutropenia and thus could be a contributing factor. Similar observations were recently described by Logue et al. with real life use of Ide-Cel^22^. Of note, incidence of infection was lower in our series, as it was the incidence of severe infection, in contrast with other published series^22,62^.

Our results are consistent with previous observations in MM patients treated with CAR-T cells^24^, as well as with the CAR-HEMATOTOX model developed by Rejeski *et al*. for predicting hematologic toxicity in patients with R/R large B cell lymphoma^25^, where baseline cytopenia and inflammatory markers correlated with duration of cytopenia. This could be associated to poor bone marrow reserve due to prior treatments, although to date no association between number of treatments and cytopenia has been found^17,18,21,22,24^. Interestingly, no association between CRS and ICANs severity and cytopenia was found in Rejeski’s work^25^, nor in our study. In contrast, Juluri *et al*. recently showed that severe CRS is associated with hematologic toxicity following CD19 CAR-T cell therapy^21^. In their study, severity of cytopenia was comparable to that seen in our patients, but its duration was substantially shorter, with a median time to ANC and platelet recovery of 8.7 days and 36.5 days respectively. Furthermore, evaluation of serum cytokines related with CRS revealed that higher IL-6 peak levels were associated with cytopenia at day 28, whereas high serum concentrations of TGF-β were associated with improved hematopoietic recovery in their work^21^. Additional biomarkers of inflammation, such as ferritin were also associated with the frequency and severity of hematological toxicity in our study. Therefore, although acute inflammation after CAR-T cell infusion, and CRS development along with lymphodepletion account partially for initial cytopenia, sustained inflammation might play an important role in the delayed hematopoietic recovery observed in some patients.

The relationship between inflammation and HSPC differentiation is well-known and has been analyzed in the context of acute syndromes, such as sepsis and hemophagocytic lymphohystiocitosis, which has also been observed as a toxicity of CAR-T cell treatment and is the paradigm of how inflammation halts hematopoiesis^13,14^. Moreover, chronic inflammation has been shown to produce HSPC exhaustion and functional damage^63,64^. Similarly, sustained inflammation caused by cytokines released by activated CAR-T cells in the bone marrow in MM patients could contribute to impaired hematopoietic recovery. These results suggest that cytokines released by activated CAR-T cells and the inflammatory environment created are at least partially responsible for the delayed hematopoietic recovery observed.

Additional biomarkers of inflammation, such as ferritin were also associated to the frequency and severity of hematological toxicity in our study. Therefore, although acute inflammation after CAR-T cell infusion, and CRS development along with lymphodepletion account partially for initial cytopenia, sustained inflammation might play an important role in the delayed hematopoietic recovery observed in some patients. The relationship between inflammation and HSPC differentiation is well-known and has been analyzed in the context of acute syndromes, such as sepsis and hemophagocytic lymphohystiocitosis, which has also been observed as a toxicity of CAR-T cell treatment and it is the paradigm of how inflammation halts hematopoiesis^13,14^. Moreover, chronic inflammation has been shown to produce HSPC exhaustion and functional damage^63,64^. Similarly, sustained inflammation caused by cytokines released by activated CAR-T cells in the bone marrow in MM patients could contribute to impaired hematopoietic recovery.

To date, there is limited knowledge regarding the underlying mechanisms of inflammation-mediated hematological toxicity after CAR-T therapy. Our *ex vivo* studies point towards a cytokine-mediated molecular rewiring of hematopoietic differentiation as a mechanism contributing to halt myeloid differentiation. As expected, characterization of supernatants from activated CAR-T cells revealed high levels of CRS-related cytokines, which could be implicated in the differentiation delay observed, as they play well-known roles in the regulation of inflammatory reactions (i.e. IL-6 and IL-10), of hematopoietic stem and progenitor cells (i.e. IL-6 and GM-CSF), and of proliferation and differentiation of B and T lymphocytes (i.e. IL-2, IL-4 and IL-15), controlling the perpetuation of the inflammatory cascade^65^. High IFNγ levels are also deleterious for HSPCs and downstream differentiation, and sustained levels of this cytokine have been shown to lead to anemia and bone marrow failure. TNFα affects myeloid precursors in a similar manner.

The results of the scRNA-seq studies also provide molecular explanation regarding regulation of cell differentiation. An immature phenotype can be explained by the expression of transcription factors involved in the initial stages of hematopoiesis, HSPC development and lineage commitment, such as *GATA2*^65,66^. This suggests that substances present in activated CAR-T supernatant create an environment that is not favorable to hematopoietic differentiation, and therefore, cells would remain in an immature quiescent state through overexpression of these TF.

From a clinical standpoint, our results suggest that early and effective control of the inflammatory cascade produced by CAR-T activation may be important to reduce hematological toxicity. Tocilizumab is currently the frontline treatment for CRS, but by blocking IL-6 receptors it results in higher circulating IL-6 levels; this might explain why higher tocilizumab use correlated with long-lasting cytopenia, both in our work and others^67^. Further studies are needed to assess whether other approaches that are based on cytokine blockage might control CRS without negatively impacting hematopoietic recovery (e.g.: anakinra, steroids, anti-TNF)^68^. Overall, our clinical findings add to the growing body of data that highlights that persistent cytopenia is a frequent and long-lasting toxicity that follows BCMA CAR-T cell therapy. It corroborates that duration of cytopenia correlates with baseline cytopenia and high peak inflammatory markers, as was already shown for CD19 directed CAR-T cells. Also, the *ex-vivo* findings indicate that activated CAR-T cells secrete substances that impair hematopoietic differentiation and reshape transcriptional programs. Thus, these data would contribute to explain the persistence of the hematopoietic toxicity observed, as damage seems to happen early in the hematopoietic hierarchy. These findings, that need to be confirmed by further studies, constitute the first mechanistic explanation for the cytopenia observed in patients treated with BCMA directed CAR-T cells.

## Data Availability

All data needed to evaluate the conclusions in the paper are present in the paper and/or the Supplementary Materials. The scRNA-seq data generated in this study have been deposited in the GEO database (GSE250444).

## ACKNOWLEDGEMENTS

We particularly acknowledge the patients and healthy donors for their participation in this study, and the Biobank of the University of Navarra for its collaboration.

## AUTHORSHIP CONTRIBUTIONS

M.L.P-B: formal analysis, investigation and writing original draft. P.R-M: formal analysis, investigation and writing original draft. M.E.C-C: formal analysis and software. N.B: formal analysis and investigation. A.Z: investigation. L.B: investigation. D.A: investigation. P.SM-U: investigation. A.V-Z: investigation. S.R-D: investigation. S.I: investigation. A.LD-C: investigation. S.H: resources and investigation. E.T: resources and investigation. J.R: resources and investigation. A.A-P: resources and investigation. J.J.L: Conceptualization. B.P: Conceptualization and resources. M.H: Conceptualization, formal analysis, funding acquisition and writing original draft. P.R-O: Conceptualization, formal analysis and writing original draft. J.S-M: Conceptualization. T.E: Conceptualization, investigation, formal analysis, funding acquisition and writing original draft. J.R.R-M: Conceptualization, formal analysis, funding acquisition and writing original draft. F.P: Conceptualization, formal analysis, funding acquisition and writing original draft.

## REFERENCES

1. June CH, Sadelain M. Chimeric antigen receptor therapy. New England Journal of Medicine. 2018;379(1):64–73.

2. Raje N, Berdeja J, Lin Y, et al. Anti-BCMA CAR T-Cell Therapy bb2121 in Relapsed or Refractory Multiple Myeloma. New England Journal of Medicine. 2019;380(18):1726–1737.

3. Martin T, Usmani SZ, Berdeja JG, et al. Ciltacabtagene Autoleucel, an Anti-B-cell Maturation Antigen Chimeric Antigen Receptor T-Cell Therapy, for Relapsed/Refractory Multiple Myeloma: CARTITUDE-1 2-Year Follow-Up. Journal of Clinical Oncology. 2022;

4. Gandhi UH, Cornell RF, Lakshman A, et al. Outcomes of patients with multiple myeloma refractory to CD38-targeted monoclonal antibody therapy. Leukemia. 2019;

5. Mikhael J. Treatment Options for Triple-class Refractory Multiple Myeloma. Clin Lymphoma Myeloma Leuk. 2020;

6. Rodriguez-Otero P, Ailawadhi S, Arnulf B, et al. Ide-cel or Standard Regimens in Relapsed and Refractory Multiple Myeloma. New England Journal of Medicine. 2023;388(11):1002–1014.

7. San-Miguel J, Dhakal B, Yong K, et al. Cilta-cel or Standard Care in Lenalidomide-Refractory Multiple Myeloma. New England Journal of Medicine. 2023;

8. Munshi NC, Anderson LD, Shah N, et al. Idecabtagene Vicleucel in Relapsed and Refractory Multiple Myeloma. N Engl J Med. 2021;384(8):705–716.

9. Berdeja JG, Madduri D, Usmani SZ, et al. Ciltacabtagene autoleucel, a B-cell maturation antigen-directed chimeric antigen receptor T-cell therapy in patients with relapsed or refractory multiple myeloma (CARTITUDE-1): a phase 1b/2 open-label study. The Lancet. 2021;

10. Gogishvili T, Danhof S, Prommersberger S, et al. SLAMF7-CAR T cells eliminate myeloma and confer selective fratricide of SLAMF7+ normal lymphocytes. Blood. 2017;130(26):2838–2847.

11. Morris EC, Neelapu SS, Giavridis T, Sadelain M. Cytokine release syndrome and associated neurotoxicity in cancer immunotherapy. Nat Rev Immunol. 2022;

12. Brudno JN, Kochenderfer JN. Recent advances in CAR T-cell toxicity: Mechanisms, manifestations and management. Blood Rev. 2019;

13. Fajgenbaum DC, June CH. Cytokine Storm. New England Journal of Medicine. 2020;

14. Chou CK, Turtle CJ. Insight into mechanisms associated with cytokine release syndrome and neurotoxicity after CD19 CAR-T cell immunotherapy. Bone Marrow Transplant. 2019;

15. Hayden PJ, Roddie C, Bader P, et al. Management of adults and children receiving CAR T-cell therapy: 2021 best practice recommendations of the European Society for Blood and Marrow Transplantation (EBMT) and the Joint Accreditation Committee of ISCT and EBMT (JACIE) and the European Haematol. Annals of Oncology. 2022;

16. Sheth VS, Gauthier J. Taming the beast: CRS and ICANS after CAR T-cell therapy for ALL. Bone Marrow Transplant. 2021;

17. Wudhikarn K, Perales MA. Infectious complications, immune reconstitution, and infection prophylaxis after CD19 chimeric antigen receptor T-cell therapy. Bone Marrow Transplant. 2022;

18. Strati P, Varma A, Adkins S, et al. Hematopoietic recovery and immune reconstitution after axicabtagene ciloleucel in patients with large B-cell lymphoma. Haematologica. 2021;

19. Neelapu SS, Locke FL, Bartlett NL, et al. Axicabtagene Ciloleucel CAR T-Cell Therapy in Refractory Large B-Cell Lymphoma. New England Journal of Medicine. 2017;

20. Locke FL, Ghobadi A, Jacobson CA, et al. Long-term safety and activity of axicabtagene ciloleucel in refractory large B-cell lymphoma (ZUMA-1): a single-arm, multicentre, phase 1–2 trial. Lancet Oncol. 2019;

21. Juluri KR, Wu QV, Voutsinas J, et al. Severe cytokine release syndrome is associated with hematologic toxicity following CD19 CAR T-cell therapy. Blood Adv. 2022;

22. Logue JM, Peres LC, Hashmi H, et al. Early cytopenias and infections after standard of care idecabtagene vicleucel in relapsed or refractory multiple myeloma. Blood Adv. 2022;

23. Fried S, Avigdor A, Bielorai B, et al. Early and late hematologic toxicity following CD19 CAR-T cells. Bone Marrow Transplant. 2019;

24. Li H, Zhao L, Sun Z, et al. Prolonged hematological toxicity in patients receiving BCMA/CD19 CAR-T-cell therapy for relapsed or refractory multiple myeloma. Front Immunol. 2022;

25. Rejeski K, Perez A, Sesques P, et al. CAR-HEMATOTOX: a model for CAR T-cell–related hematologic toxicity in relapsed/refractory large B-cell lymphoma. Blood. 2021;

26. Rodriguez-Marquez P, Calleja-Cervantes ME, Serrano G, et al. CAR Density Influences Antitumoral Efficacy of BCMA CAR T cells and Correlates with Clinical Outcome. Sci Adv. 2022;2022.01.19.22269515.

27. Berastegui N, Ainciburu M, Romero JP, et al. The transcription factor DDIT3 is a potential driver of dyserythropoiesis in myelodysplastic syndromes. Nat Commun. 2022;

28. Peng J, Serrano G, Traniello IM, et al. SimiC enables the inference of complex gene regulatory dynamics across cell phenotypes. Commun Biol. 2022;

29. Van Oekelen O, Aleman A, Upadhyaya B, et al. Neurocognitive and hypokinetic movement disorder with features of parkinsonism after BCMA-targeting CAR-T cell therapy. Nat Med. 2021;

30. Shah N, Chari A, Scott E, Mezzi K, Usmani SZ. B-cell maturation antigen (BCMA) in multiple myeloma: rationale for targeting and current therapeutic approaches. Leukemia. 2020;

31. Uhlen M, Karlsson MJ, Zhong W, et al. A genome-wide transcriptomic analysis of protein-coding genes in human blood cells. Science (1979). 2019;

32. Karlsson M, Zhang C, Méar L, et al. A single–cell type transcriptomics map of human tissues. Sci Adv. 2021;

33. Stelzer G, Rosen N, Plaschkes I, et al. The GeneCards suite: From gene data mining to disease genome sequence analyses. Curr Protoc Bioinformatics. 2016;

34. Hay SB, Ferchen K, Chetal K, Grimes HL, Salomonis N. The Human Cell Atlas bone marrow single-cell interactive web portal. Exp Hematol. 2018;

35. DePasquale EAK, Schnell D, Dexheimer P, et al. cellHarmony: cell-level matching and holistic comparison of single-cell transcriptomes. Nucleic Acids Res. 2019;

36. Silvestre-Roig C, Hidalgo A, Soehnlein O. Neutrophil heterogeneity: Implications for homeostasis and pathogenesis. Blood. 2016;

37. Yang P, Li Y, Xie Y, Liu Y. Different faces for different places: Heterogeneity of neutrophil phenotype and function. J Immunol Res. 2019;

38. Grieshaber-Bouyer R, Nigrovic PA. Neutrophil heterogeneity as therapeutic opportunity in immune-mediated disease. Front Immunol. 2019;

39. Garg B, Mehta HM, Wang B, et al. Inducible expression of a disease-associated ELANE mutation impairs granulocytic differentiation, without eliciting an unfolded protein response. Journal of Biological Chemistry. 2020;

40. Xie X, Shi Q, Wu P, et al. Single-cell transcriptome profiling reveals neutrophil heterogeneity in homeostasis and infection. Nat Immunol. 2020;

41. Ichikawa M, Yoshimi A, Nakagawa M, et al. A role for RUNX1 in hematopoiesis and myeloid leukemia. Int J Hematol. 2013;

42. Hayashi Y, Harada Y, Harada H. Myeloid neoplasms and clonal hematopoiesis from the RUNX1 perspective. Leukemia. 2022;

43. Katsumura KR, Bresnick EH. The GATA factor revolution in hematology. Blood. 2017;

44. You X, Zhou Y, Chang YI, et al. Gata2 19.5 enhancer regulates adult hematopoietic stem cell self-renewal and T-cell development. Blood Adv. 2022;

45. Tsai FY, Orkin SH. Transcription factor GATA-2 is required for proliferation/survival of early hematopoietic cells and mast cell formation, but not for erythroid and myeloid terminal differentiation. Blood. 1997;

46. De Filippo K, Rankin SM. CXCR4, the master regulator of neutrophil trafficking in homeostasis and disease. Eur J Clin Invest. 2018;

47. Eash KJ, Means JM, White DW, Link DC. CXCR4 is a key regulator of neutrophil release from the bone marrow under basal and stress granulopoiesis conditions. Blood. 2009;

48. Avellino R, Delwel R. Expression and regulation of C/EBPα in normal myelopoiesis and in malignant transformation. Blood. 2017;

49. Zamani F, Shahneh FZ, Aghebati-Maleki L, Baradaran B. Induction of CD14 expression and differentiation to monocytes or mature macrophages in promyelocytic cell lines: New approach. Adv Pharm Bull. 2013;

50. Jakubzick C V., Randolph GJ, Henson PM. Monocyte differentiation and antigen-presenting functions. Nat Rev Immunol. 2017;

51. Brand M, Ranish JA. Proteomic/transcriptomic analysis of erythropoiesis. Curr Opin Hematol. 2021;

52. Tusi BK, Wolock SL, Weinreb C, et al. Population snapshots predict early haematopoietic and erythroid hierarchies. Nature. 2018;

53. Verma R, Su S, McCrann DJ, et al. RHEX, a novel regulator of human erythroid progenitor cell expansion and erythroblast development. Journal of Experimental Medicine. 2014;

54. Lord KA, Abdollahi A, Hoffman-Liebermann B, Liebermann DA. Proto-oncogenes of the fos/jun family of transcription factors are positive regulators of myeloid differentiation. Mol Cell Biol. 1993;

55. Buitenhuis M, Van Deutekom HWM, Verhagen LP, et al. Differential regulation of granulopoiesis by the basic helix-loop-helix transcriptional inhibitors Id1 and Id2. Blood. 2005;

56. Evrard M, Kwok IWH, Chong SZ, et al. Developmental Analysis of Bone Marrow Neutrophils Reveals Populations Specialized in Expansion, Trafficking, and Effector Functions. Immunity. 2018;

57. Khoyratty TE, Ai Z, Ballesteros I, et al. Distinct transcription factor networks control neutrophil-driven inflammation. Nat Immunol. 2021;

58. Huber R, Pietsch D, Panterodt T, Brand K. Regulation of C/EBPβ and resulting functions in cells of the monocytic lineage. Cell Signal. 2012;

59. Kelly LM, Englmeier U, Lafon I, Sieweke MH, Graf T. MafB is an inducer of monocytic differentiation. EMBO Journal. 2000;

60. Schüler A, Schwieger M, Engelmann A, et al. The MADS transcription factor Mef2c is a pivotal modulator of myeloid cell fate. Blood. 2008;

61. Roy B, Bhattacharjee A, Xu B, et al. IL-13 signal transduction in human monocytes: phosphorylation of receptor components, association with Jaks, and phosphorylation/activation of Stats. J Leukoc Biol. 2002;

62. Kambhampati S, Sheng Y, Huang CY, et al. Infectious complications in patients with relapsed refractory multiple myeloma after BCMA CAR T-cell therapy. Blood Adv. 2022;

63. Clapes T, Lefkopoulos S, Trompouki E. Stress and non-stress roles of inflammatory signals during HSC emergence and maintenance. Front Immunol. 2016;

64. Bousounis P, Bergo V, Trompouki E. Inflammation, aging and hematopoiesis: A complex relationship. Cells. 2021;

65. Ye F, Huang W, Guo G. Studying hematopoiesis using single-cell technologies. J Hematol Oncol. 2017;

66. Vicente C, Conchillo A, García-Sánchez MA, Odero MD. The role of the GATA2 transcription factor in normal and malignant hematopoiesis. Crit Rev Oncol Hematol. 2012;

67. Tie R, Li H, Cai S, et al. Interleukin-6 signaling regulates hematopoietic stem cell emergence. Exp Mol Med. 2019;51(10):.

68. Zhang L, Wang S, Xu J, et al. Etanercept as a new therapeutic option for cytokine release syndrome following chimeric antigen receptor T cell therapy. Exp Hematol Oncol. 2021;10(1):4– 7.

