## Supplemental material for "Molecular mechanisms promoting long-term cytopenia after BCMA CAR-T therapy in Multiple Myeloma"

**Supplementary figures**

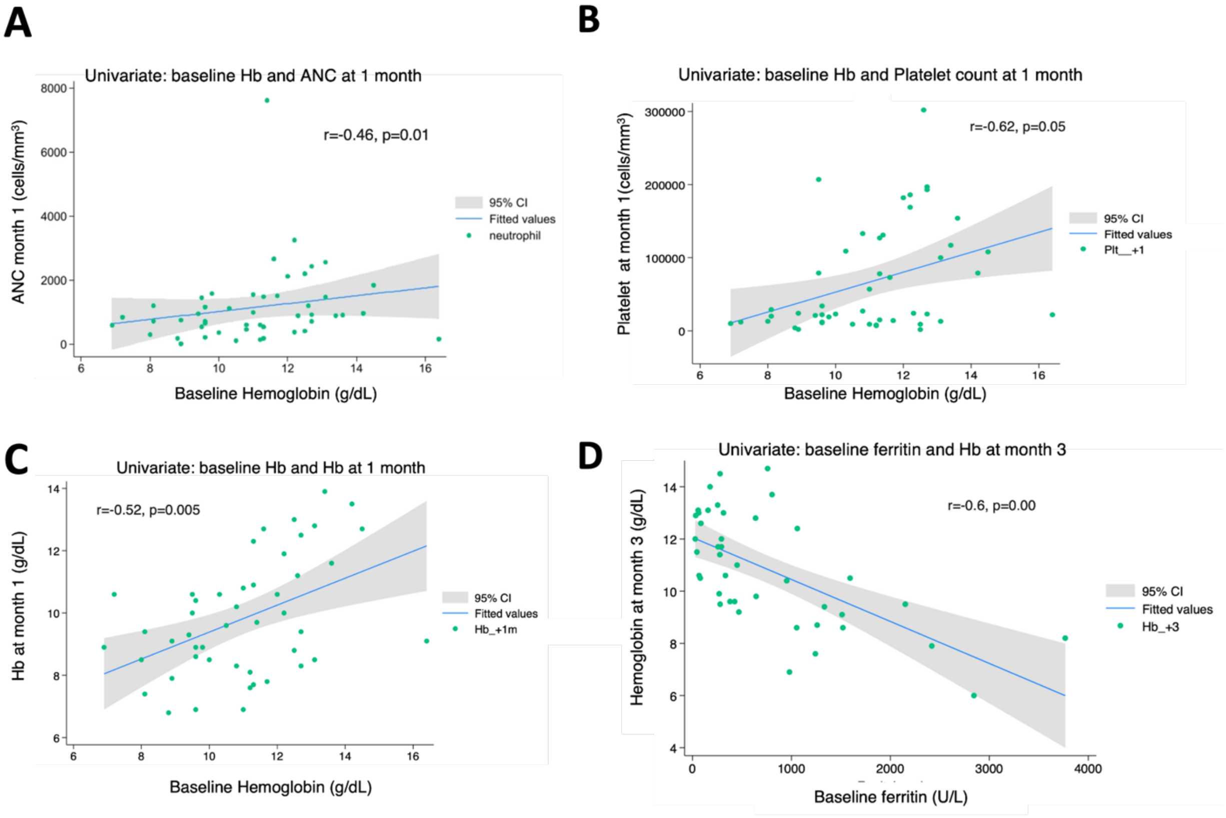


**Figure S1. Baseline cytopenia (defined as the blood counts before lymphoapheresis) correlate significantly with the presence of long term cytopenias.** Univariate analysis of the influence of baseline hemoglobin with blood counts: baseline hemoglobin correlates significantly with neutrophil **(A)**, platelet **(B)** and hemoglobin (Hb) **(C)** counts one month after infusion. Baseline inflammatory markers such as ferritin **(D)** also correlate with long lasting cytopenia, such as anemia at month 3, as depicted in panel D. The Spearman correlation coefficient (r) and respective P values are provided. A positive r value indicates a positive correlation, and a negative r value indicates a negative correlation. Light grey shading indicates the 95% confidence bands of the best-fit line from the simple linear regression.

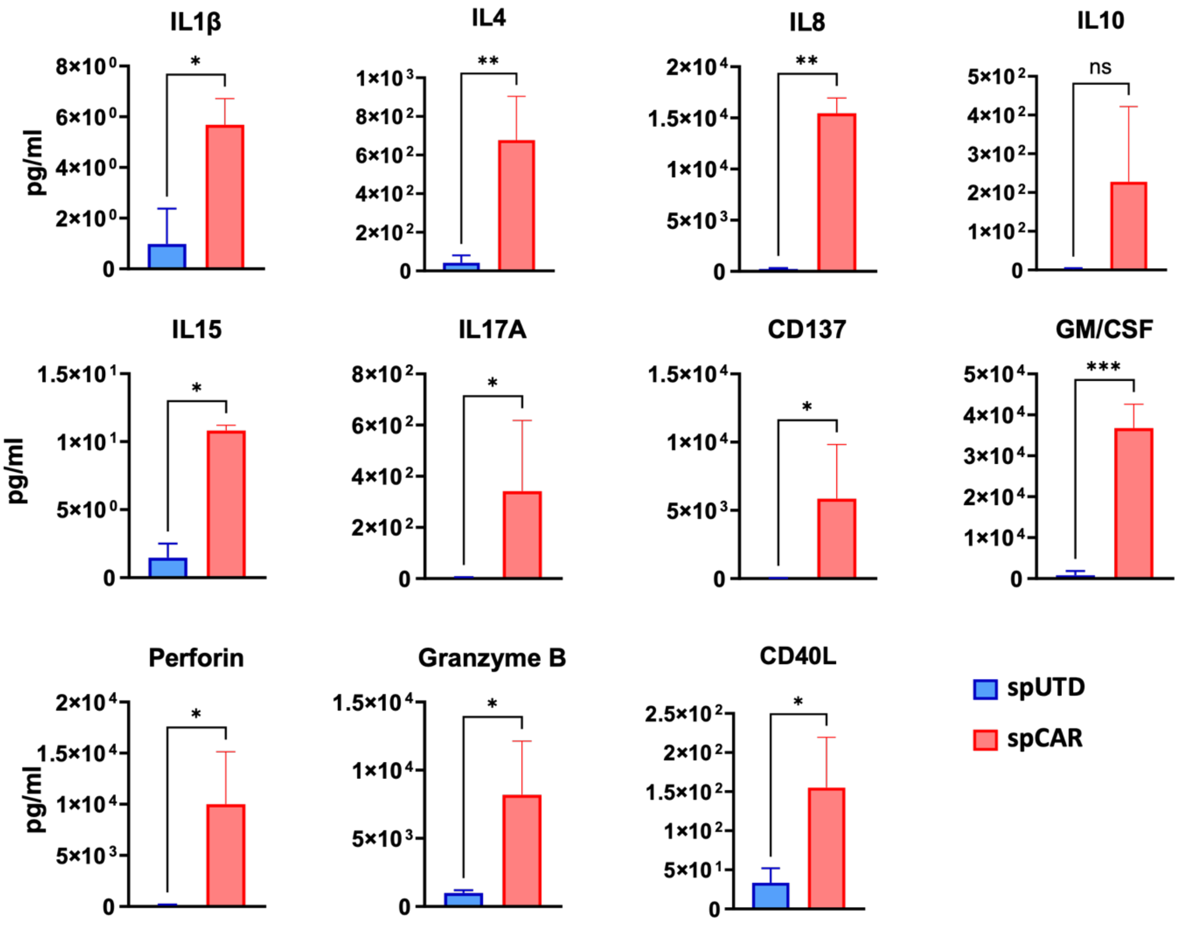


**Figure S2. Concentration of cytokines was significantly higher in the spCAR.** Cytokines present in the supernatant of activated BCMA CAR-T cells (red) and in the supernatant of control untransduced lymphocytes (blue) after 48h of co-culture with MM cell line U266 at 1:1 effector:target ratio. A custom 15-plex ProcartaPlex panel was performed, including IL1β, IL4, IL8, IL10, IL15, IL17A, CD137, GM/CSF, perforin, granzyme B, CD40L, and cytokines from Fig.2. The concentration of the pro-inflammatory cytokines analyzed was significantly higher in the spCAR when compared to its control counterpart, that presented negligible concentrations of the cytokines studied. Welch's test. *$p<0.05$; **$p>0.01$; ***$p>0.001$.

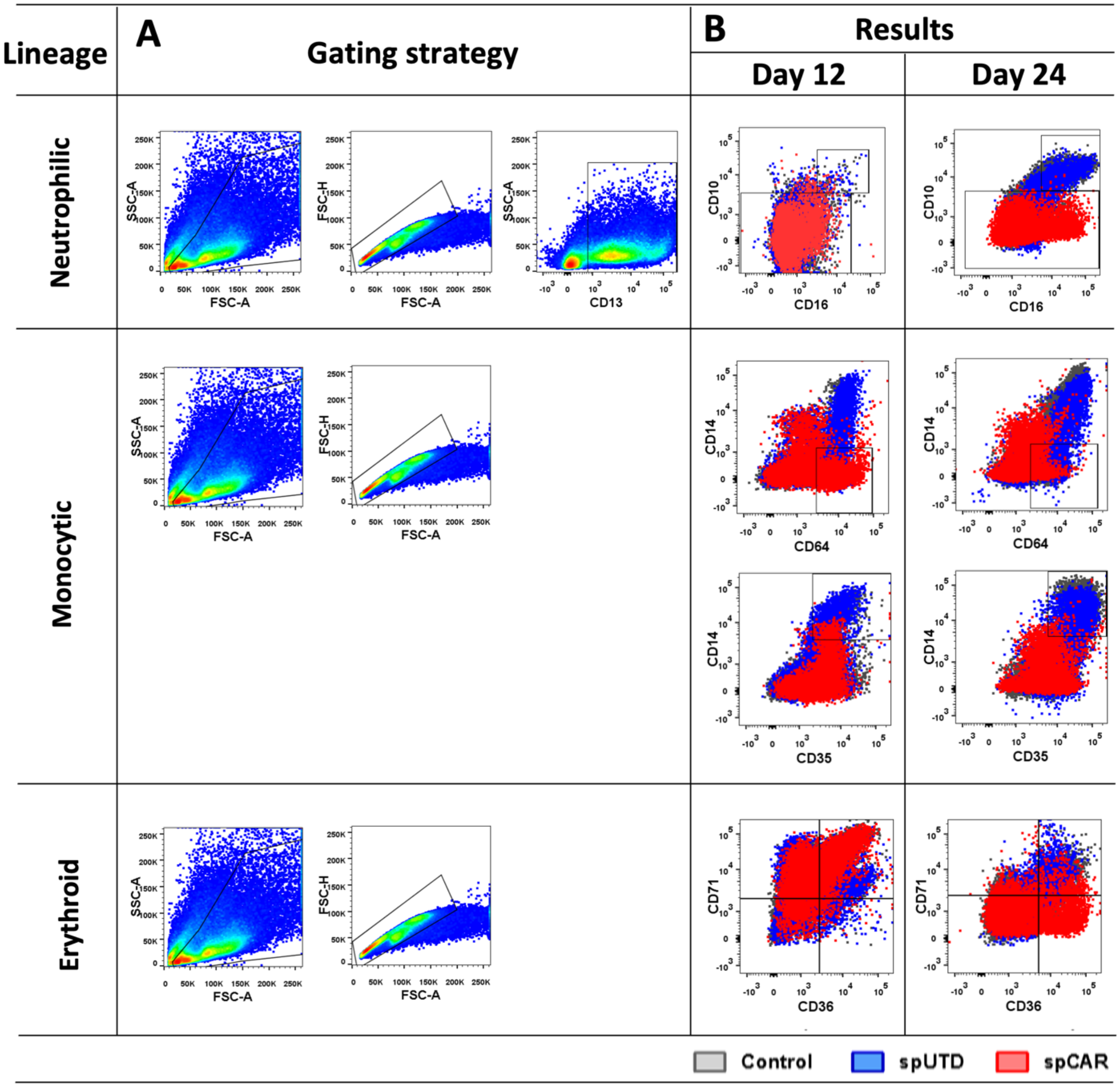


**Figure S3. HSPCs differentiated with the supernatant of activated CAR-T cells presented less mature phenotypes. (A)** FACS gating strategy. After selection of cells (SSC-A, FSC-A) and singlets (FSC-H, FSC-A), population markers for each lineage were studied. Neutrophilic lineage also was selected for $CD13^{+}$ cells previously. **(B)** FACS gating results of HSPCs differentiated in the control (gray), spUTD (blue) and spCAR (red) conditions at day 12 and 24 of differentiation. Gates of less differentiated and more differentiated cells is shown for the three lineages, neutrophilic ($CD10^{-}$; $CD10^{+}CD16^{+}$), monocytic ($CD14^{-}CD64^{+}$; $CD14^{+}CD35^{+}$) and erythroid ($CD71^{+}CD36^{-}$; $CD71^{+}CD36^{+}$) respectively.

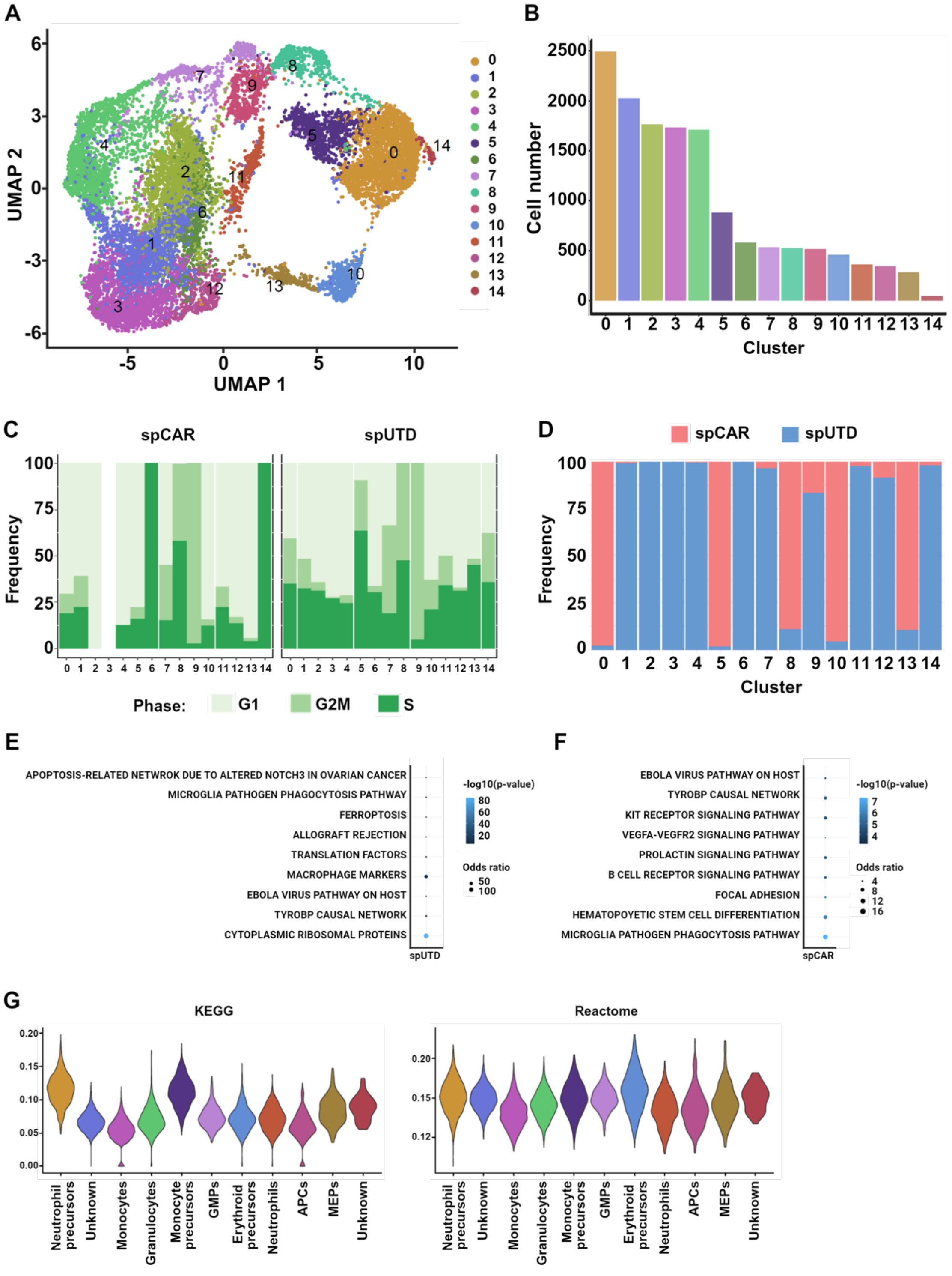


**Figure S4. Characterization of CD34$^+$ cells at single cell level.** scRNAseq of ex-vivo liquid culture differentiation samples of healthy CD34+ cells at day 24 after addition of spCAR or spUTD as control was performed. **(A)** An overview of the 14248 cells analyzed in this study. UMAP plot showing the 15 clusters that were identified. **(B)** Bar plots showing the number of cells per cluster. **(C)** Bar plot showing the degree of G1, G2M and S phases signatures in each cluster and each condition (spCAR or spUTD) **(D)** Bar plot showing the percent contribution of cells from each condition (spCAR or spUTD) to the different clusters. **(E)** Gene ontology analysis showed maturation pathways associated with cluster of monocytes, corresponding to cells exposed to spUTD. **(F)** Gene ontology analysis showed differentiated pathways associated with cluster of monocyte precursors, corresponding to cells exposed to spCAR. **(G)** Gene signature of apoptosis using KEGG and reactome.

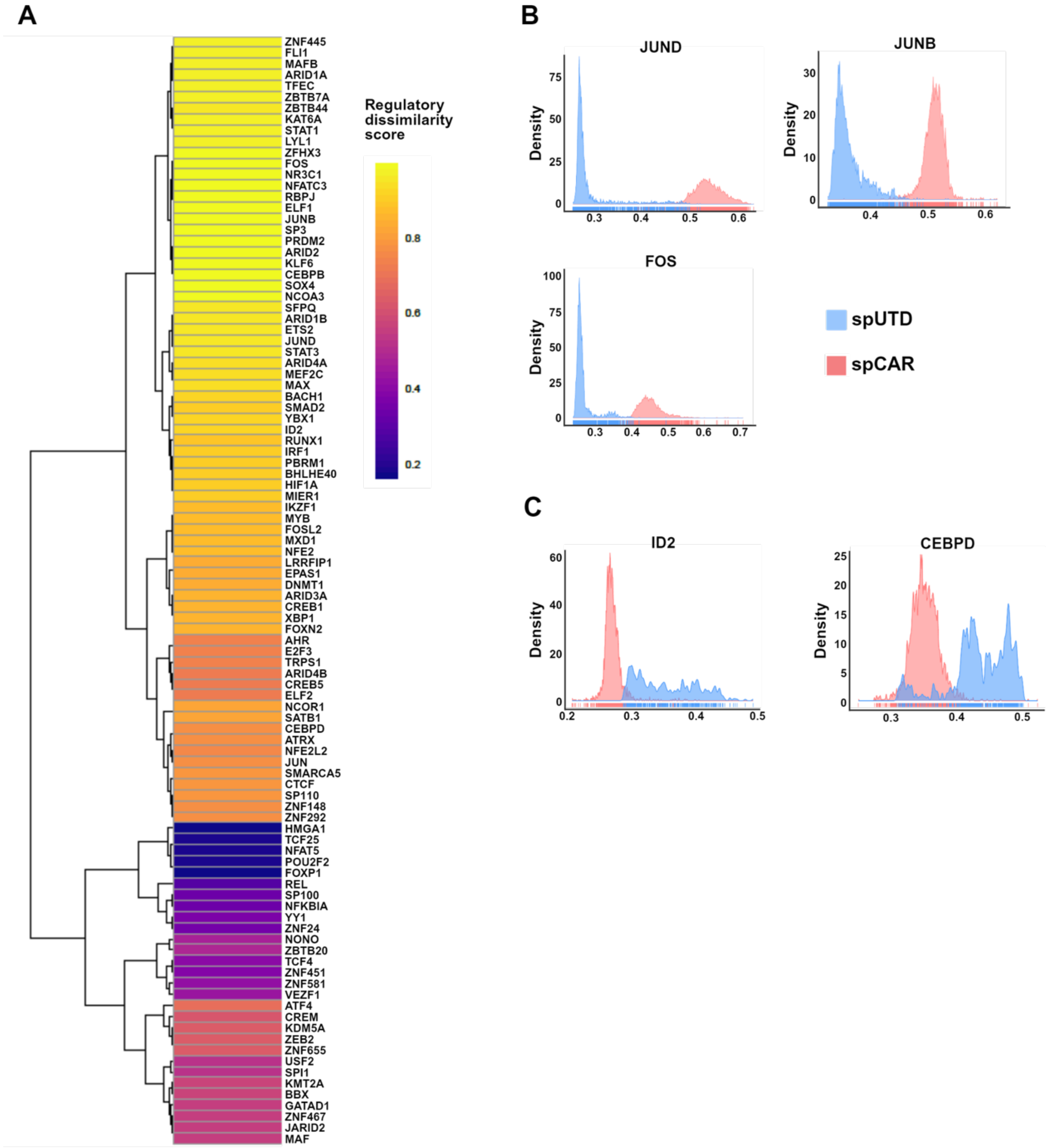


**Figure S5. Analysis of GRNs in cells differentiated in presence of spCAR or spUTD.** We applied SimiC, a novel GRN inference algorithm for scRNA-seq data, to infer GRNs associated to more mature phenotypes or still in differentiation process. **(A)** Heatmap showing the regulatory dissimilarity score between cells exposed to spUTD or spCAR of the different regulons. **(B)** Histograms showing the activity score of *JUND, JUNB,* and *FOS* regulons. **(C)** Histograms showing the activity score of *ID2* and *CEBPD* regulons.
